# What constitutes safe and effective dose titration of methadone and buprenorphine/naloxone? protocol for a population-based target trial emulation

**DOI:** 10.64898/2026.07.23.26358700

**Authors:** Momenul Haque Mondol, Michelle Zanette, Jeong Eun Min, Megan Kurz, Robert William Platt, Shaun Seaman, Paxton Bach, Mohammad Ehsanul Karim, Maria Eugenia Socías, Paul Gustafson, Jason M Sutherland, Bohdan Nosyk

**Affiliations:** School of Population and Public Health, University of British Columbia, Vancouver, Canada; Centre for Advancing Health Outcomes, Vancouver, British Columbia, Canada; Department of Statistics and Data Science, University of Barishal, Barishal, Bangladesh; Faculty of Health Sciences, Simon Fraser University; Burnaby, British Columbia, Canada; Department of Epidemiology, Biostatistics and Occupational Health, McGill University, Montreal, Quebec, Canada; MRC Biostatistics Unit, Cambridge, UK; Department of Medicine, University of British Columbia, Vancouver, British Columbia, Canada; British Columbia Centre on Substance Use, Vancouver, British Columbia, Canada; Department of Statistics, University of British Columbia, Vancouver, British Columbia, Canada; Centre for Health Services and Policy Research, University of British Columbia, Vancouver, Canada

## Abstract

**Introduction:** Clinical guidelines for managing opioid use disorder (OUD) recommend incremental dose titration for methadone and buprenorphine/naloxone to achieve a safe therapeutic maintenance dose. Dose titration recommendations are based on clinical experience, pharmacologic rationale, and expert consensus, with limited real-world evidence, especially in settings with widespread fentanyl use, where traditional titration schedules may be insufficient to address the higher opioid tolerance among people initiating treatment. This study aims to determine the comparative effectiveness of alternative titration schedules on completed induction and time to all-cause mortality.

**Methods and analysis:** We will conduct a population-based retrospective cohort study using linked data from nine provincial health administrative databases. The study population will include adults (≥18 years) in British Columbia, Canada, who initiated methadone or buprenorphine/naloxone between 01/01/2010 and 30/06/2022. Primary outcomes are completed induction (defined as no dose increase for ≥two weeks without any intervening decrease) and time to all-cause mortality, with overdose-related acute care visits or overdose-related death and treatment discontinuation as secondary outcomes. Using a target trial framework, this study will implement a ‘clone-censor-weight’ approach to estimate the per-protocol effects of sustained titration schedules capturing adherence to and deviation from clinical guidelines. Sensitivity analyses will assess the robustness of findings through cohort and timeline restrictions as well as alternative exposure and outcome definitions.

**Discussion:** This study will generate real-world evidence on the comparative effectiveness of alternative titration schedules of methadone and buprenorphine/naloxone on completed induction and all-cause mortality. The findings will support evidence-informed updates to OAT guidelines and clinical decision-making in British Columbia and other jurisdictions facing escalating opioid-related harms.

## Introduction

### Opioid agonist treatment initiation

Methadone and buprenorphine/naloxone are two forms of opioid agonist treatments (OAT) used to treat opioid use disorder (OUD), which affected 40.5 million people globally in 2017.[1] Both medications provide substantial protection against overdose risk and all-cause mortality among people with OUD (PWOUD).[2–5] In British Columbia (BC), Canada, now in the 10^th^ year of provincial public health emergency due to unregulated drug toxicity deaths, OAT retention is declining,[6,7] with drop out frequently occurring early in treatment.[8] Induction on OAT is a critical period during which the medication dose is titrated to achieve a desired therapeutic effect (i.e., eliminate withdrawal symptoms) while minimizing side effects. The risk of overdose and mortality is highest during the induction period.[5,9,10] Methadone has a long and variable half-life and can accumulate in the body over several days, necessitating incremental dose increases every few days to avoid toxic serum levels. Increasing the dose too rapidly can result in fatal overdose, while delayed increases may leave withdrawal symptoms inadequately managed, contributing to early treatment discontinuation or continued use of unregulated opioids.[11] With buprenorphine/naloxone, a high-affinity partial mu-opioid receptor agonist, rapid titration in the presence of residual full agonists may precipitate withdrawal, while insufficient or overly cautious dose increases may fail to adequately suppress withdrawal symptoms and cravings.[12] An effective titration schedule must therefore balance speed and safety to stabilize individuals, improve retention, and reduce mortality.

### Clinical guidelines on OAT dose titration

With the introduction of high-potency synthetic opioids such as fentanyl into the unregulated drug supply,[13] titration of OAT has become more challenging as opioid tolerance is generally accepted to have increased among people who are regularly exposed.[14,15] In response, prescribers across Canada and the United States have been adapting their titration practices to achieve higher doses more rapidly, outpacing both research and clinical guidelines.[16,17] More recent clinical guidelines for the management of OUD in BC emphasize the need for individually tailored titration protocols and note that traditional heroin-era titration approaches may be insufficient in the context of increasing fentanyl-driven opioid tolerance among PWOUD.[18]

Specifically, the 2017 BC guidelines recommended methadone to be titrated by 5-10mg every 5 or more days.[19] Reflecting clinical experience in the fentanyl era, the 2023 update to the guidelines revised this recommendation to 5-10 mg every 3-5 days for individuals with lower or unknown opioid tolerance. In people with higher opioid tolerance, such as those with confirmed fentanyl exposure, higher increases of up to 15 mg every 3 days is recommended, up to a total daily dose of 85 mg, followed by increases of 10 mg every 3-5 days thereafter. Also new to the 2023 update, and based primarily on clinical experience, rapid titration protocols (e.g., starting with 30-40mg, with up to 3×10mg as needed per day)[20] have been described during hospitalization in monitored settings (e.g., hospital wards, inpatient detox facilities, bed-based treatment facilities) to reduce dropout and support earlier stabilization, though careful monitoring for opioid toxicity remains essential.[21]

For buprenorphine/naloxone, BC guidelines similarly recommend individuals be initiated with 2mg/0.5mg-4mg/1mg based on withdrawal severity and increases of 2mg/0.5mg-4mg/1mg every 1-3 hours, as needed. The 2023 BC guidelines retained this approach but increased the maximum daily dose from 24mg/6mg to 32mg/8mg per day for individuals with high opioid tolerance, exceeding the Canadian product monograph maximum of 24mg/6mg. Once the risk window for precipitated withdrawal has passed, more aggressive symptom-guided titration has long been recommended. Earlier guideline suggested as reaching 12-16 mg within the first day, which is no longer reflective of current practice. In addition, low-dose induction protocols, also known as micro-dosing, which start with sub-therapeutic doses (e.g., 0.5mg-1mg doses several times daily, with small, serial increases made over anywhere from 3-10 days) while continuing full agonist opioids, have emerged as an alternative to the traditional induction.[22] These protocols eliminate the need for people to be in withdrawal prior to initiation and might be beneficial for individuals using fentanyl or at high risk of destabilization. The growing use of high and low dose induction approaches reflects a recognition that titration is not uniform and should be matched to individual clinical stability, withdrawal severity, and opioid tolerance.[18]

Clinical guidelines from other jurisdictions recommend similar titration schedules both for methadone and buprenorphine/naloxone (see **Table 1**).

**Table 1.** Clinical guidelines of titration schedules of methadone and buprenorphine/naloxone.

| **Guideline** | **Methadone** | **Buprenorphine/naloxone** |
| --- | --- | --- |
| BCCSU, BC, CAN, 2017[19] | 5-10mg every ≥5 day | 2/0.5mg-4/1mg per increase (and several times as needed), up to 16/4mg on Day 2, up to 24/6mg after. |
| BCCSU, BC, CAN, 2023[18] | *Rapid induction:* 3×10mg as needed every day; | *Low-dose induction:* ≤2/0.5mg every day before reaching 8/2mg or 16/4mg; |
|  | *Traditional induction:* 15mg max every 3 days before reaching 85mg and 10mg max every 3-5 day onwards for clients with known high tolerance; 10mg max every 3-5 day for clients with lower or unknown tolerance. | and >2/0.5mg every day after reaching 8/2mg or 16/4mg;<br>*Traditional induction:* 2/0.5mg-4/1mg per increase (up to the maximum 32/8mg). |
| META: PHI, ON, CAN, 2022[23,24] | 10-15mg every 3-5 day for clients taking <75mg daily, and 10mg every 5-7 day for clients taking ≥75mg daily. | *Low-dose induction:* 0.5/0.125mg every day until 12/3mg, and then 2/0.5mg per increase as needed. |
| CAMH, CAN, 2021[25] | 5mg every ≤5 day for clients who initiated with ≤10mg; 5-10mg every 3-5 day for clients who initiated with 5-20mg; 5-15mg every 3-5 day for clients who initiated with 5-30mg. | *Low-dose induction:* Doubling every day (0.5/0.125 is the starting dose);<br>*Traditional induction:* 2/0.5-6/1.5mg every day for older adults |
| CRISM, CAN, 2018 | AB & SK: ≤10mg every 3 day, ≤10mg every 4 day, and ≤5mg every 5 day for clients who initiated 30mg, 20mg, and 10mg respectively;<br>MB: 5-10mg every 3-4 day<br>ON & NB: 10-15mg every 3-5 day, 5-10mg every 3-5 day, and 5mg every 5 day for clients who initiated 30mg, 20mg, and 10mg respectively;<br>QC: 5-20mg every 4-6 day | AB, SK, MB, ON, and NB: 2/0.5mg-4/1mg per increase to reach daily maximum 16/4mg;<br>QC: 2/0.5-4/1mg per increase to reach daily maximum of 8/2-16/4mg. |
| ASAM, USA, 2020[26] | ≤10mg every 5 day | 2-8mg per increase |
| ASAM, USA, 2015[27] | 5-10mg every 7 day | 2-4mg per increase |
| Q-Land, AUS, 2023[28] | 5-10mg every ≥3 day | *Low-dose induction:* Gradual build from 0.4mg until reaching 8mg on day 7, then build to 12-16mg on day 12-14 |
| AUS, 2014[29] | 5-10mg every 3-5 day | 2-8mg per increase |
| NSW, AUS, 2018[30] | 5-10mg every 3-5 day | 2-4mg per increase |
| UK, 2017[31] | 5-10mg everyday (max. 30mg a week) | Doubling every day from initial 4mg. |

**Legend:** ASAM, American Society of Addiction Medicine; AUS, Australia; BC, British Columbia; AB, Alberta; SK, Saskatchewan; MB, Manitoba; ON, Ontario; NB, New Brunswick; QC, Quebec; BCCSU, British Columbia Centre on Substance Use; CAN, Canada; CAMH, Center for Addiction and Mental Health; Q-Land, Queensland state; CRISM, Canadian Research Initiative on Substance Misuse; NSW, New South Wales; UK, the United Kingdom; USA, the United States of America.

For methadone, recommendations range from 5-15mg increases every 3-5 days.[19,25,28–34] For buprenorphine/naloxone, guidelines from Ontario[23,24] and Australia[29] also recommend low-dose induction, and other jurisdictions only advise titration schedules of 2/0.5mg-4/1mg or 2/0.5mg-8/2mg per increase.[19,25,28,31–34]

Prescribing practices during the COVID-19 pandemic across Canada,[35,36] the United States,[37,38] Australia,[39] New Zealand,[40] and the United Kingdom[41] were also shaped by broader changes in expanded take-home dosing, telehealth-based care, and reduced urine drug screening to maintain treatment continuity under limited in-person contact. For titration specifically, less frequent in-person monitoring may have altered the pace of dose increase, although the direction and magnitude of any such effect remain unclear.

### Evidence of effectiveness of titration schedules

Evidence regarding the comparative effectiveness of alternative titration schedules on treatment retention, risk of overdose and mortality is sparse. Notably, dose titration guidance in BC was developed through expert consensus due to a lack of evidence regarding titration of OAT during the fentanyl era.[18,19] Some studies signal more rapid titration may be associated with improved treatment retention,[42–45] with qualitative reports indicating a preference for rapid titration among people with high tolerance or prior experience with OAT.[24,46,47]

#### Impact on effectiveness

A recently published retrospective cohort study of individuals titrating on methadone in the United States (2020-2023) found higher doses on day 7 of treatment, whether reflecting higher starting doses, faster titration, or both, were associated with higher rates of retention at 30 days post initiation.[43] Notably, the study potentially selected for participants with less OAT experience and more stability as it excluded individuals who initiated methadone with >30mg or missed a dose in the first week. Similarly, a retrospective cohort study of 13,560 methadone recipients in Ontario, Canada (2017-2022) found that receiving an initial dose increase 4-6 days after treatment initiation was associated with a substantially lower hazard of treatment discontinuation during the first week of follow-up (weighted hazard ratio: 0.55, 95% compatibility interval 0.51-0.60), compared with remaining on the initial dose through day 6.[44] However, individuals had to remain in treatment long enough to receive a dose increase between days 4 and 6 in this study, potentially introducing immortal time bias.[48] In addition, distinct titration strategies were not evaluated, limiting applicability of findings within real-world methadone titration regimens. Further, a secondary analysis of the OPTIMA randomized control trial (RCT), including 167 participants with prescription opioid type OUD across Canada (including fentanyl; 2017-2020),[45] found higher methadone dose ranges and longer titration durations were associated with increased retention at 24 weeks, and that faster titration rates were associated with reduced opioid use during the maintenance phase.[45]

For buprenorphine/naloxone, no association was found between the duration and rate of titration with retention or opioid use.[45] However, these findings were limited to prescription opioid type OUD and excluded people who primarily used street drugs (other than fentanyl). Another study specifically investigated buprenorphine titration trajectories among PWOUD randomized to receive methadone or buprenorphine at 8 opioid treatment programs in the United States as part of the START trial.[42] Titration trajectories were derived from collected data and were not randomized. They found that participants with higher baseline withdrawal scores and faster titration were less likely to drop out in the first 7 days of treatment compared to those who titrated more slowly; however, they found no differences in retention at 28 days. Since buprenorphine dosing was witnessed daily among this sample, findings may not be reflective of settings in which take-home dosing is permitted during titration.

#### Impact on safety

The literature regarding safety outcomes such as overdose and mortality during methadone titration is largely restricted to small sample retrospective cohort studies and case reports, with a strong focus on rapid titration protocols. A retrospective cohort study from Maryland, USA, found that among 25 hospitalized patients with OUD who were rapidly titrated on methadone, there were no reports of overdoses or mortality in hospital and at 30-days post-discharge.[49] Similarly, a retrospective cohort study among 98 inpatients in Vancouver, BC, found rapid titration was well-tolerated with few adverse events (i.e. administration of naloxone for sedation and ICU transfer for observation, observed in 1.2% of their sample).[21] In San Francisco, a case series of 17 patients who received rapid titration reported four (21%) experienced mild to moderate sedation events. [50] In contrast, an older case series of 10 deaths in Victoria, Australia, described patients who died a mean of three days after commencing methadone maintenance from general practitioners, with a mean starting dose of 53mg. The authors emphasized the risks of relatively high starting doses in individuals without demonstrated opioid tolerance. Another forensic review of 206 methadone-associated deaths in Victoria, Australia (2001–2005) found that 25% occurred within 14 days of commencing opioid replacement therapy, with a median starting dose of 35 mg and frequent dose increases of up to 25 mg per day. The authors attributed these induction-period deaths largely to iatrogenic toxicity from excessive starting doses and rapid dose escalation.[51] However, it is important to note that this evidence pertains to monitored inpatient settings with close clinical oversight, and these findings may not apply to community settings. For buprenorphine, the aforementioned START trial found no differences in adverse events (e.g., persistent headache, non-cardiac chest pain, suicidal ideation, overdose) among individuals on different titration trajectories.[42] A 2021 systematic review focused on low-dose induction of buprenorphine and the authors found that, among 19 case studies/series and 1 feasibility study, most studies did not report precipitated withdrawal, and could not report on comparative effectiveness or safety due to the lack of studies comparing low-dose titration to standard dosing.[52] However, a 2025 systematic review found 44 articles on low-dose induction, indicating a significant surge of the use of this strategy in clinical practice.[53] The authors found minimal or mild withdrawal symptoms are commonly reported despite the intention of this strategy in avoiding withdrawal, though the literature is limited to case reports/series and single-arm observational studies.

#### Clients’ perspectives on titration

A qualitative analysis of 29 participants from the NAOMI clinical trial, which compared supervised injectable diacetylmorphine or hydromorphone to oral methadone in Canada, found that those receiving methadone identified the possibility of rapid titration as one of methadone’s positive attributes.[46] A narrative systematic review of 47 qualitative studies published between 2005 and 2023, spanning community and criminal justice settings across multiple countries, found that withdrawal during induction and a lack of collaborative decision-making were common.[54] Similarly, among 30 methadone patients in rural Vermont and New Hampshire[55] almost all reported daily withdrawal during induction and used fentanyl to manage symptoms rather than for recreation, in one participant’s words, “not to get high, but just to get by.” Participants’ three key recommendations were faster induction, expanded take-home access, and broader split dosing availability.

### Objective

In summary, much of the evidence on titration schedules for both methadone and buprenorphine lack comparator groups to determine differences in effectiveness between emerging (such as rapid induction of methadone and low-dose or high-dose induction of buprenorphine/naloxone) and traditional titration strategies.[21,49,50,52] Limited sample sizes[21,49,50] and heterogeneity in reporting, outcome measures, individual and program characteristics and follow-up periods[42,43,45,49] among titration strategies limit the ability to synthesize the current state of literature and draw definitive conclusions. Real-world evidence is needed to inform clinical guidelines and support collaborative clinical decision-making among prescribers and clients. Therefore, the objective of this study is to compare the effectiveness of alternative titration schedules of methadone and buprenorphine/naloxone on completed induction and time to all-cause mortality as observed in clinical practice in BC from 2010 to 2022.

## Materials and Methods

### Study design

We propose a population-based retrospective cohort study aiming to emulate target trials (i.e., hypothetical RCTs) for individuals initiating methadone and buprenorphine/naloxone, respectively, between 01/01/2010, and 30/06/2022, in BC, Canada. Although RCTs are the gold standard for determining comparative effectiveness, they are often time-intensive and may lack generalizability to the broader population of PWOUD.[56] By leveraging large-scale health administrative datasets and advanced statistical techniques, observational studies can address many common biases and yield treatment effect estimates based on real-world evidence.[57–60] Key elements of the target trials and their emulations are summarized in **Table 2**.

**Table 2.** Key components of the emulated target trials to assess titration strategy using observational data.

| Protocol components | Hypothetical target trial | Emulation of the trial from BC health administrative data |
| --- | --- | --- |
| Eligibility criteria | We will include adults (age ≥18 years), initiated MET or BNX between 01/01/2010, and 30/06/2022. Individuals are not incarcerated, pregnant, and have no cancer, or palliative care history. | Same.<br>Incident user design (no history of OAT dispensation, dating back to 01/01/1996) and prevalent new-user design (repeated attempts of OAT after a dispensation gap of ≥5 days for methadone, ≥6 days for buprenorphine/naloxone) will be considered. |
| Treatment strategies | For MET trial, four titration strategies will be defined: Dose increases of (i) up to 10 mg every 5 or more days, (ii) up to 10 mg every 3 or more days, (iii) up to 15 mg every 3 or more days, and (iv) up to 20 mg every 3 or more days. In strategies (iii) and (iv), dose increases are restricted to 10 mg or lower once the daily dose reaches 85 mg or higher.<br>For BNX trial, titration strategies will be defined: (i) up to 4 mg every 2 or more days, (ii) up to 2 mg every 1 or more | Same. |
|  | days, (iii) up to 4 mg every 1 or more days, and (iv) up to 8 mg every 1 or more day. Dose increases will be restricted to 4 mg or lower once the daily dose reaches 8 mg or higher for the strategy (ii) and 16 mg or higher for strategy (iv).[18] |  |
| Assignment procedures | Randomization. Prescribers, healthcare teams, and individuals are aware of the treatment status. | Each individual has a clone created for each titration strategy of target trials of MET and BNX. Prescribers, healthcare teams, and individuals are aware of the treatment status. |
| Time zero | The date of randomization | The date of OAT initiation |
| Outcomes | An indicator of completed induction (defined as no dose increases in $\geq 14$ days without any intervening decrease), and time to all-cause mortality, with time to overdose-related acute care visit or death and OAT discontinuation (no dispensation for $\geq 5$ days for MET, $\geq 6$ days for BNX) as secondary outcomes. | Same. Due to the lack of inpatient information on OAT dispensation in our linked database, we will assume OAT receipt prior to hospitalization is continued daily with no dose change throughout the duration of hospitalization. |
| Follow-up | From time zero to the earliest date of primary outcome of interest, deviation from the assigned titration strategy, treatment switch, taper initiation, pregnancy, incarceration, initiation of cancer or palliative care, death, six months after time zero or 12/31/2022 (see **Table 3**). | Same. |
| Causal contrasts | Per-protocol effect | Same. |
| Analysis plan | Cumulative incidence (risk) difference will be estimated using inverse probability weighted pooled logistic regression model. Weights are the function of baseline and post-baseline covariates (updated daily) to control for the selection bias induced by censoring. | Same as target trial, except for the potential immortal time bias will be addressed using a clone-censor-weight approach.[61] |

**Legend:** BC, British Columbia; MET, methadone; BNX, buprenorphine/naloxone; OAT, opioid agonist treatment

We will use a data linkage of nine provincial health administrative databases in BC, where residents are required to participate in a single-payer health insurance system. In BC, methadone and buprenorphine/naloxone may be prescribed in office-based settings and are dispensed at community pharmacies, with take-home doses authorized by the prescriber based on assessments of clinical stability. Using the BC PharmaNet database,[62] which captures all community pharmacy dispensations, we will identify dispensation dates and doses (refer to **S1 Table** for a list of drug/product identification numbers) to define OAT episodes. Additional databases include the Discharge Abstract Database[63] (DAD, documenting hospitalizations), Medical Services Plan[64] (MSP, containing physician billing records), BC Vital Statistics [65] (BCVS, capturing deaths and underlying causes), BC Provincial Corrections [66] (recording incarceration entries and releases), Perinatal Care Database[67] (covering maternal and infant outcomes), National Ambulatory Care Reporting System[68] (NACRS, documenting Emergency Department visits), Client Roster[69] (providing demographic and geographic data), and the BC Social Development and Poverty Reduction database[70] (tracking social assistance). These datasets are linked at the individual level using a de-identified personal health number assigned by the BC Ministry of Health.[71] A description of these databases and the timeline for data extraction is detailed in **S2 Table**.

### Study population

Our study population will include adults (≥18 years) who initiated OAT with methadone or buprenorphine/naloxone between 01/01/2010 and 30/06/2022, allowing a minimum of 6 months of potential follow-up through 31/12/2022. Individuals with no current incarceration, no known pregnancy, and no cancer care or palliative care history at time zero will be included. Pregnant individuals will be excluded due to the acceleration of maternal metabolism during pregnancy which may influence dosing requirements,[18,72,73] while those incarcerated will be excluded due to the distinct treatment protocols followed during incarceration.[74] Time zero is the date of first dispensation of methadone or buprenorphine/naloxone. We will divide the total dispensed dose evenly across the days supplied to calculate daily exposure to each medication. Due to a lack of inpatient dispensation data in our linked database, OAT receipt prior to hospitalization will be assumed to continue daily at an unchanged dose throughout the hospital stay.[19] OAT episodes are defined as continuous receipt of OAT dispensations without interruptions of ≥5 days for methadone and ≥6 days for buprenorphine/naloxone (see **S1 Fig.**). These thresholds align with BC guidelines recommending a return to starting doses after these durations.[18] Episodes preceded by hospitalization will also be excluded to reduce potential misclassification of titration pace (described in **Exposure** section). To capture clinical experiences in both first-time and repeated attempts at treatment, we will use incident user (no OAT dispensation since 01/01/1996) and prevalent new-user designs (no OAT dispensation in past 30 days).[16,75–77]

### Exposure

Four titration strategies are defined for each of the methadone and buprenorphine/naloxone trials, reflecting a gradient from the most conservative (reference) to the most aggressive approach.[18,19] Each strategy is characterized by two dimensions: maximum allowable dose increment and minimum permissible interval between successive dose increases. For the methadone trial, strategies are: (i) dose increases of up to 10 mg at intervals of 5 or more days (reference); (ii) up to 10 mg at intervals of 3 or more days; (iii) up to 15 mg at intervals of 3 or more days; and (iv) up to 20 mg at intervals of 3 or more days. For strategies (iii) and (iv), once the daily dose reached 85 mg or higher, the maximum allowable increment is restricted to 10 mg or less, consistent with 2023 BC guidelines recommending heightened caution at higher methadone doses.[18]

For the buprenorphine/naloxone trial (doses refer to the buprenorphine component), we consider the following dose titration strategies: (i) dose increases of up to 4 mg every 2 or more days (reference); (ii) up to 2 mg every 1 or more days; (iii) up to 4 mg every 1 or more days; and (iv) up to 8 mg every 1 or more days. For strategy (ii), dose increases were restricted to 4 mg or lower once the daily dose reached 8 mg or higher; for strategy (iv), this restriction applied from 16 mg onward. [19]

Notably, these strategies lack a lower threshold on the magnitude of each dose increase and an upper threshold on the number of days permitted between increases. These strategies thus reflect the maximum permissible pace for a given strategy. The implications of these definitional choices will be explored in several sensitivity analyses.

### Outcome

Primary outcomes will include completed induction and time to all-cause mortality. Completed induction is a binary outcome, defined as reaching a minimum of 14 days of continuous OAT without dose changes within 6 months of treatment initiation. Since the in-hospital dose is imputed rather than observed (see **Study population** section), hospital days do not count toward the 14-day no-dose-change window, which is suspended on admission and resumes on discharge, so that completed induction reflects observed community dosing only. Time to OAT discontinuation (defined as no dispensation for ≥5 days for methadone, ≥6 days for buprenorphine/naloxone) and overdose-related acute care visits (identified using ICD-10 codes T40, T42.4, and T43.6) or overdose-related death will be considered as secondary outcomes.

### Study follow-up

Since the titration schedule is not observable at time zero (i.e., each OAT episode is compatible with all four titration strategies at time zero), we will implement a ‘clone-censor-weight’ (CCW) approach to emulate per-protocol target trials.[78,79] The hypothetical target trial is a randomized trial of usual OAT care with a single intervention; a ceiling on the pace and magnitude of dose escalation defined by assigned titration strategy. Individuals would be randomized to one of four disjoint strategies that differ only in this ceiling; care would otherwise proceed as usual. Each eligible treatment episode will be replicated into four clones, one assigned to each titration strategy. All clones carry identical baseline characteristics, ensuring complete initial balance across titration strategies (analogous to randomization in RCTs). A clone will be artificially censored at the first date on which the observed dosing pattern becomes inconsistent with the assigned strategy (e.g., dose increase exceeds the maximum allowable increment or increase earlier than the minimum allowable interval) (see **Fig. 1**).

**Fig 1.**
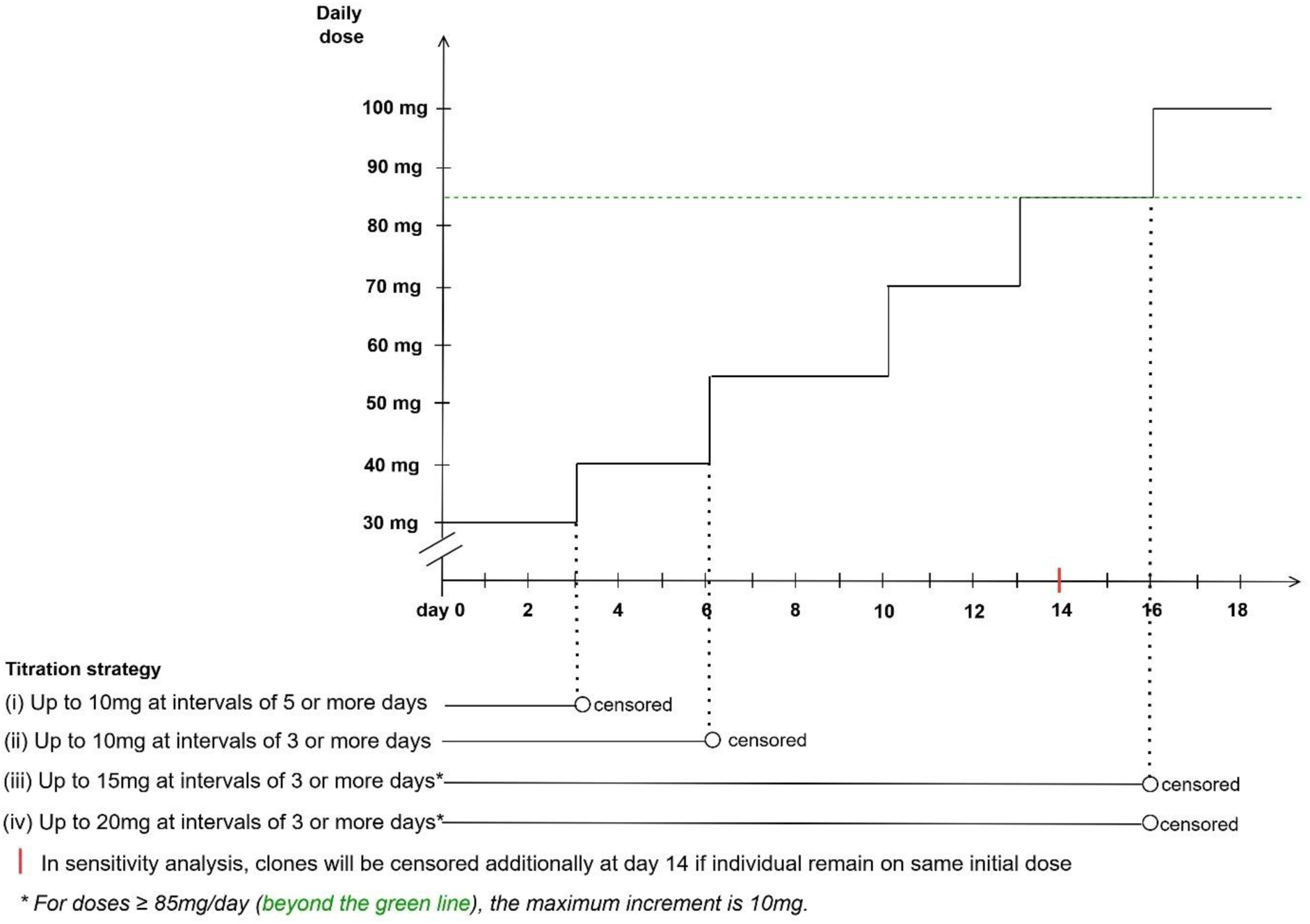
Censoring process for a hypothetical individual receiving methadone Legend: We generate four clones of this hypothetical individual receiving methadone, each assigned to one titration strategy. Each clone is censored at the first time it deviates from its assigned strategy. Clone (i) is censored at day 3 due to an increase in dose occurring earlier than the minimum allowed interval under its assigned strategy (indicated by a dotted vertical line). Clone (ii) is censored at day 6 due to a dose increase of 15mg, exceeding the maximum permitted increment under its assigned strategy. Clones (iii) and (iv) are censored at day 16 due to a dose increase of 15 mg, which exceeds the maximum allowed increment of 10 mg at doses ≥85 mg/day. Since the strategies are ordered from most to least restrictive, these censoring times are non-decreasing across the clones (days 3, 6, 16 and 16, respectively): a clone still uncensored under a stricter strategy is necessarily still uncensored under the more permissive ones. This nesting of the clones’ at-risk time is an expected feature of the cloning and does not reflect overlap between the treatment arms.

For each clone, follow-up begins at time zero, is measured in daily intervals, and ends at the earliest date of the primary outcome event or an end of follow-up event. End of follow-up events are: six months after time zero or 12/31/2022 (C1); pregnancy, incarceration, initiation of cancer or palliative care, treatment switch or taper initiation (defined as two consecutive daily dose reductions without an intervening increase) (C2); artificial censoring due to deviation from the assigned titration strategy (C3). For the completed-induction analysis, which evaluates a binary outcome within six months of time zero, death and OAT discontinuation are treated as failure (outcome indicator is assigned as zero). The structure of censoring events by outcome is detailed in **Table 3**.

**Table 3.**
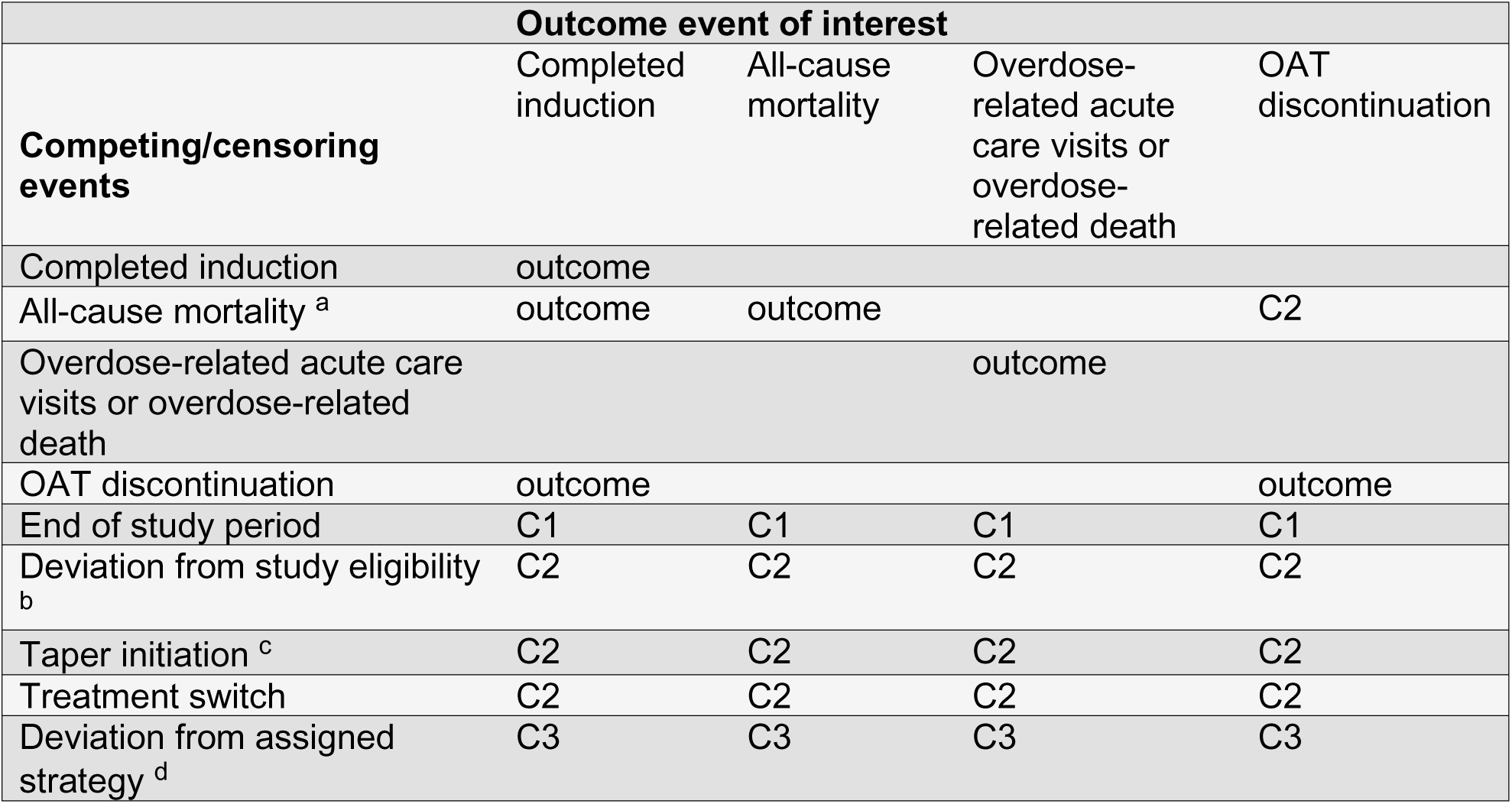
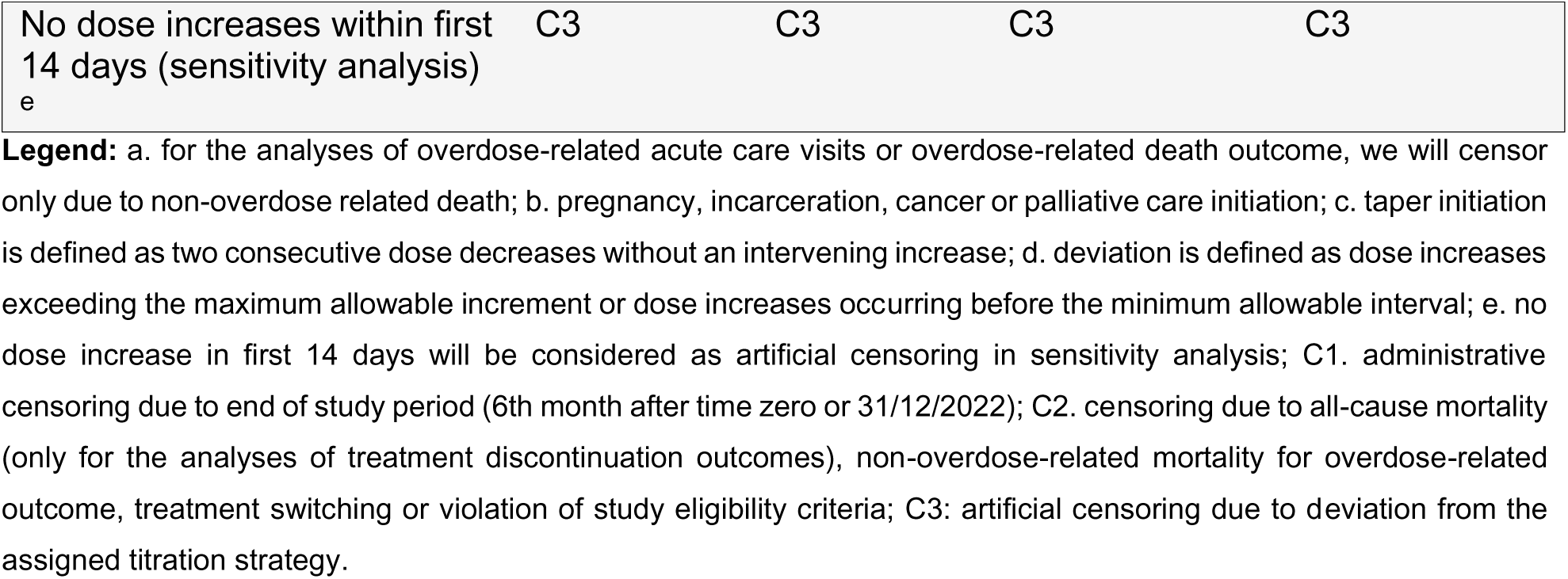
Censoring indication in the primary and secondary analyses for comparing titration strategies of methadone and buprenorphine/naloxone.

| Competing/censoring events | Outcome event of interest |  |  |  |
| --- | --- | --- | --- | --- |
|  | Completed induction | All-cause mortality | Overdose-related acute care visits or overdose-related death | OAT discontinuation |
| Completed induction | outcome |  |  |  |
| All-cause mortality <sup>a</sup> | outcome | outcome |  | C2 |
| Overdose-related acute care visits or overdose-related death |  |  | outcome |  |
| OAT discontinuation | outcome |  |  | outcome |
| End of study period | C1 | C1 | C1 | C1 |
| Deviation from study eligibility <sup>b</sup> | C2 | C2 | C2 | C2 |
| Taper initiation <sup>c</sup> | C2 | C2 | C2 | C2 |
| Treatment switch | C2 | C2 | C2 | C2 |
| Deviation from assigned strategy <sup>d</sup> | C3 | C3 | C3 | C3 |
| No dose increases within first 14 days (sensitivity analysis)<br>e | C3 | C3 | C3 | C3 |

**Legend:** a. for the analyses of overdose-related acute care visits or overdose-related death outcome, we will censor only due to non-overdose related death; b. pregnancy, incarceration, cancer or palliative care initiation; c. taper initiation is defined as two consecutive dose decreases without an intervening increase; d. deviation is defined as dose increases exceeding the maximum allowable increment or dose increases occurring before the minimum allowable interval; e. no dose increase in first 14 days will be considered as artificial censoring in sensitivity analysis; C1. administrative censoring due to end of study period (6th month after time zero or 31/12/2022); C2. censoring due to all-cause mortality (only for the analyses of treatment discontinuation outcomes), non-overdose-related mortality for overdose-related outcome, treatment switching or violation of study eligibility criteria; C3: artificial censoring due to deviation from the assigned titration strategy.

The six-month window is chosen so that follow-up is long enough for completed induction to be observed in all clones that achieve it, while remaining short enough to link titration strategies to proximal outcomes.[44]

### Covariate selection

We will control for a comprehensive set of time-fixed and time-varying covariates identified through a systematic review,[13,80,81] applying a modified disjunctive cause criterion[82] to guide covariate selection. These covariates include socio-demographic factors, measures of comorbidity, OAT-related and prescriber and system-level factors. Socio-demographic factors will include age, sex, rural versus urban region, unstable housing in the past five years, receipt of income assistance in the past year, incarceration in the past year, and calendar year of treatment initiation. Measures of comorbidity will include the Charlson Comorbidity Index, as well as indications of severe mental disorder, alcohol use disorder, hepatitis C virus infection, asthma, chronic obstructive pulmonary disease, and chronic pain, many defined with validated case-finding algorithms through hospitalization, outpatient care and drug dispensation records and updated daily. Treatment-related factors will include the dose at OAT initiation, history of overdose-related events in the past year, dispensation of non-OAT opioids (including prescription opioids prior to OUD diagnosis), dispensation of psychiatric or sedative medications in the past month, urine drug testing in the past week, treated tobacco use disorder, and attachment to the OAT initiator (>50% physician billing in the past 12 months is attributed to the prescriber who initiated OAT). Among prevalent new users we will also control for cumulative duration of prior OAT dispensations, time since last OAT dispensation, and history of completed induction. Prescriber and system-level factors will include prescriber reimbursement model (alternative payment plan versus fee-for-service), practice size (large >300 clients, medium 16-300, small ≤15), OAT delivery for methadone, receipt of a prior opioid prescription, and receipt of virtual care visits for telephone or video consultations. Calendar year will be included to capture changes in the unregulated drug supply and other temporal changes that may affect clinical practice and health outcomes (e.g., the COVID-19 pandemic) (see **Table 4**).

**Table 4.** Time fixed and time-varying covariates to estimate the inverse probability of censoring weights.

| Variable | Category | How the variable is defined and measured |
| --- | --- | --- |
| *Socio-demographic factors* |  |  |
| Age | Continuous | Based on birth date information from the client registry. |
| Sex | Male/Female | Based on medical records that record sex. |
| Living in rural region | Yes/No | Based on the population of the assigned local health area[83] |
| Homelessness/unstable housing in past 5 years<sup>‡</sup> | Past year, Past 1-5 years, Never/>5 years | Defined through ICD10: Z59.0, Z59.1 from physician billing or hospital discharge records or 3 consecutive records of no fixed address from SDPR |
| Incarcerated in past year<sup>‡</sup> | Yes/No | Admission to BC correctional facilities and identified through records in the past year from the provincial corrections database. |
| Receipt of income assistance in the past year<sup>‡</sup> | Yes/No | Identified through records in SDPR and PharmaCare plan information on medication dispensations, using records within the past year. |
| Calendar year of time zero | 2010 to 2022 | Determined based on the dispensation date from PharmaNet. |
| *Comorbidity* |  |  |
| Severe mental disorder (ever)<sup>‡, a</sup> | Yes/No | Includes major depressive disorder, bipolar disorder, and schizophrenia, and identified through ICD codes if diagnosed ever and updated if diagnosed during episode follow-up. |
| Alcohol use disorder (ever)<sup>‡, a</sup> | Yes/No | Identified through ICD codes and DIN/PIN, if diagnosed ever and updated if diagnosed during episode follow-up. |
| Hepatitis C virus (ever)<sup>‡, a</sup> | Yes/No | Identified through ICD codes if diagnosed ever and updated if diagnosed during episode follow-up. |
| Asthma (ever)<sup>‡, a</sup> | Yes/No | Identified through ICD codes* and DIN/PIN within the past year, updated during the follow-up. |
| Chronic pain in past year<sup>‡, a</sup> | Yes/No | Identified through ICD codes within the past year, updated during follow-up. |
| Chronic obstructive pulmonary disease<sup>‡, a</sup> | Yes/No | Identified through ICD codes within the past year, updated during the follow-up. |
| Charlson comorbidity index in past year<sup>‡</sup> | 0 and above | Using the Charlson comorbidity index[84] and hospitalization records within the past year, and updated during the follow-up. |
| Dispensations of psychiatric or sedative medication in the past month<sup>‡, a</sup> | Yes/No | Identified through DIN/PIN of dispensations in the past 30. Updated during the follow-up. |
| *Factors related to OAT experience* |  |  |
| Dose at OAT initiation | Continuous | Identified through dispensations in PharmaNet.[85] |
| Duration of OAT dispensation in past year | Continuous | Identified through dispensations in PharmaNet.[85] |
| History of completed induction of OAT in past year | Yes/No | Identified through dispensations in PharmaNet.[85] |
| History of overdose-related events in past year | Yes/No | Identified through ICD codes within the past year. |
| Dispensations opioid other than OAT in the past month<sup>‡, a</sup> | Yes/No | Identified through DIN/PIN and a case-finding algorithm used prior[86] and updated during the follow-up. |
| Received prescription opioids other than OAT prior to OUD diagnosis<sup>a</sup> | Yes/No | Identified through DIN/PIN and a case-finding algorithm used prior[86] and updated during the follow-up. |
| Treated tobacco use disorder in past year<sup>‡, a</sup> | Yes/No | Based on ICD-10 codes and indication of nicotine replacement therapy in PharmaNet |
| Attachment to the prescriber<sup>‡, a</sup> | Yes/No | Identified based on billing history from the physician. More than 50% of physician billing attributed to the prescriber in the past 12 months |
| Funding of prescribers who initiated OAT: alternative payment plan | Yes/No | Alternative payment plan indicated by the absence of a medical services plan billing record (capturing fee-for-service billing) at initial OAT dispensation. |
| Urine drug test in the past week<sup>‡</sup> | Yes/No | Receipt of urine drug test from physician billing records |
| Practice size<sup>‡</sup> | Large, medium, small | Determined by network analysis; number of unique OAT clients in past 12months. Large for >300 clients, medium for 16-300 clients and small for ≤15 clients. |
| *COVID-19 related factors* |  |  |
| Virtual care<sup>‡</sup> | Yes/No | Virtual care indicated in physician billing records.[87,88] |
| OAT delivery for methadone<sup>‡</sup> | Yes/No | Indication of delivery of methadone in PharmaNet |
| Receipt of prescriber safer supply (PSS)<sup>‡</sup> | Yes/No | Prior PSS dispensations in PharmaNet.[85] |

**Legend:** ICD: International Classification of Diseases; SDPR: Social Development and Poverty Reduction; DIN: drug identification number; PIN: product identification number; <sup>‡</sup> Time dependent variable will be updated daily; a. ICD codes for each condition and DIN/PINs for different medications can be found elsewhere.[13]

### Statistical analysis

#### Inverse probability of censoring weighting

We will use inverse probability of censoring weights (IPCW) to account for time-varying confounding by indication (i.e., selection bias) induced by censoring during follow-up under the CCW framework. Separate analyses will be conducted for methadone and buprenorphine/naloxone. For each trial, we will fit pooled logistic regression models[89] to estimate the probability of remaining uncensored at each follow-up day under C2 and C3 censoring mechanisms as described earlier (see **Table 3**). Each model will include baseline covariates, time-varying covariates lagged by one day to preserve temporal ordering between covariate assessment and censoring events, and time from day 0 (modelled flexibly using restricted cubic splines). For each censoring mechanism, day-specific stabilized weights will be estimated as:[90]

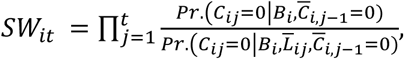

where *SW_it_* is the stabilized weight for the *i*th clone at time *t*, *C_ij_* is the censoring status for the *i*th clone at *j*th time, *C_ij_* is the censoring history for the *i*th clone at *j*th time, *B* is the list of time-fixed covariates, *L_ij_* is the time-dependent covariate history for the *i*th clone at *j*th time and Pr. denotes predicted probabilities from the pooled logistic regression models. The numerator model (conditioning on baseline covariates only) stabilises the weights and reduces variance inflation.[91] The final analytic weight for each clone-day observation will be calculated as the product of the stabilized weights across the two censoring mechanisms. We will assess the performance of censoring models:[92,93] by examining the distribution of stabilized weights over follow-up; by verifying that the stabilised weights have a mean of approximately one, as a departure would indicate model misspecification; and by examining artificial censoring status against covariate values across strategies and follow-up time to identify any subgroups with a near-zero proportion remaining uncensored, indicating violations of the positivity assumption.[94] If extreme weights are observed, weights will be truncated at the 99th percentile. Primary analyses will use truncated weights, and sensitivity analyses will assess the impact of using untruncated weights.[95]

#### Outcome models

For completed induction outcome within six months, we will fit weighted logistic regression models using the final analytic weights. Outcome models will include indicators for titration strategy and baseline covariates. However, the time to all-cause mortality will be analyzed as a discrete-time event using weighted pooled logistic regression,[89] including indicators for titration strategy, baseline covariates, flexible functions of follow-up time using restricted cubic splines, and interaction terms between strategy and time to allow treatment effects to vary over follow-up. From these models, we will estimate 6-month cumulative incidence under each titration strategy using regression standardization. For each strategy, predicted probabilities from the weighted outcome model will be averaged across the study population. To construct two-sided 95% compatibility (‘confidence’) intervals,[95] we will use parametric bootstrap approach based on the asymptotic distribution of the model parameters.[96] After fitting the weighted pooled logistic model, we will draw 500 sets of regression coefficients from a multivariate normal distribution with mean equal to the estimated coefficients and variance equal to the estimated robust sandwich covariance matrix.[61] For each simulated parameter set, cumulative incidences will be calculated using the method described above. Compatibility intervals will be obtained from the empirical distribution of the estimates.[96]

### Subgroup and sensitivity analyses

To assess the robustness of our primary findings, we will conduct a series of prespecified subgroup and sensitivity analyses, interpreted as exploratory and hypothesis-generating rather than as bases for definitive conclusions. For subgroup analyses, we will stratify by initial dose (as defined according to a prior comparative effectiveness study[97]), and clinically relevant client characteristics, including prior treatment disengagement, duration of previous OAT, completed induction in past year, sedative co-prescription in past month, chronic pain in past year, mental health comorbidities, and care setting. We will also examine populations of particular policy relevance, including adolescents, individuals incarcerated in past year, and those initiated OAT after 04/01/2016 when BC declared a public health emergency in response to rising fentanyl-related overdose deaths.[13] For sensitivity analyses, we will apply alternative definitions of titration strategy (no dose change as the reference group), completed induction (reaching at 60mg of methadone or 16 mg of buprenorphine/naloxone with the 2-weeks thresholds of no dose increase; 3-weeks threshold of no dose increase), OAT discontinuation (14- and 30-day gaps), and prevalent new users (no OAT dispensation in past 30 days), and extended follow-up from 6 to 12 months. We will additionally redefine censoring events during follow-up (no dose increase during the first 14 days will cause artificial censoring). Proposed subgroup and sensitivity analyses are summarized in **Table 5**. Cumulative incidence difference with 95% compatibility intervals will be presented as exploratory and hypothesis generating findings using forest plots.

**Table 5.**
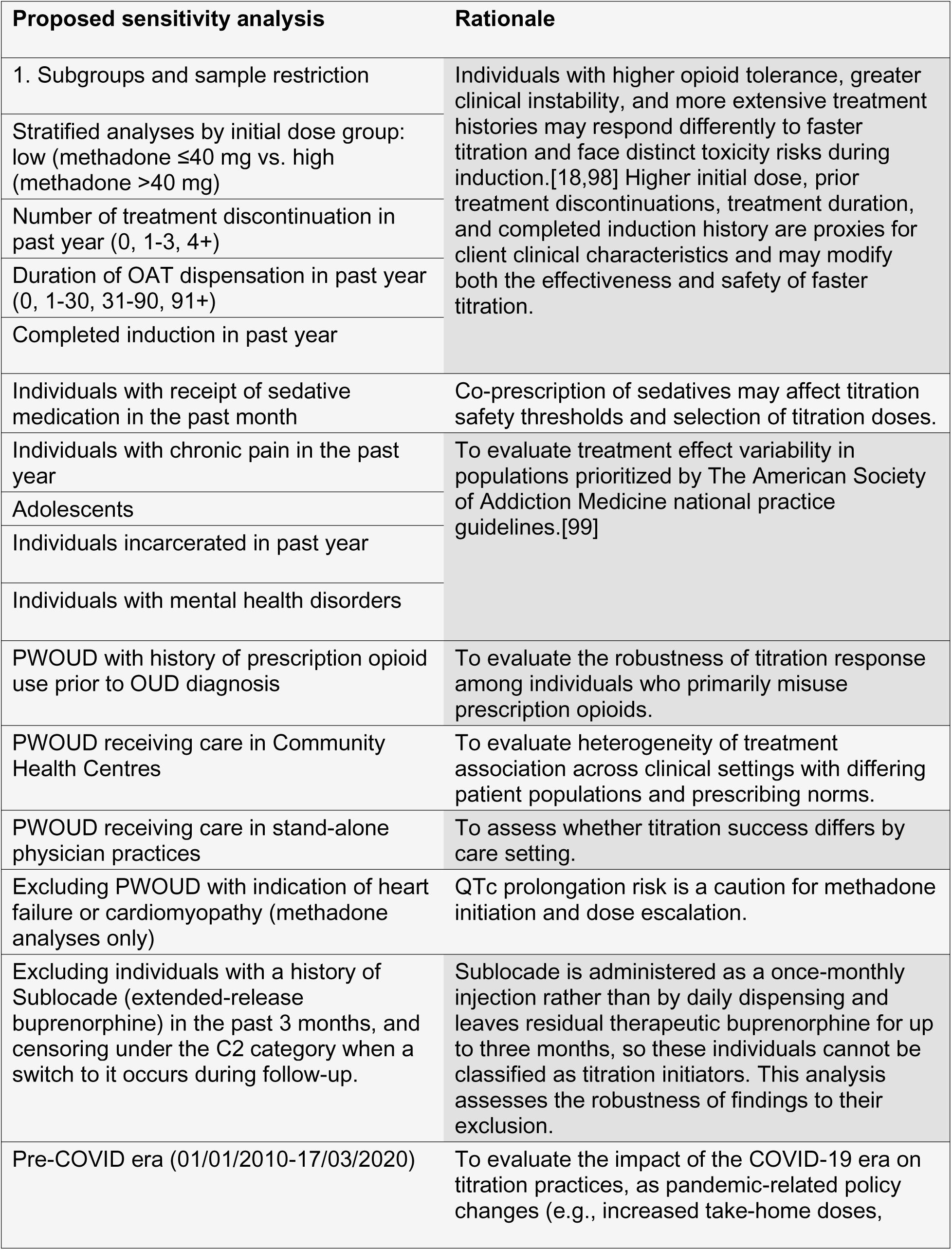
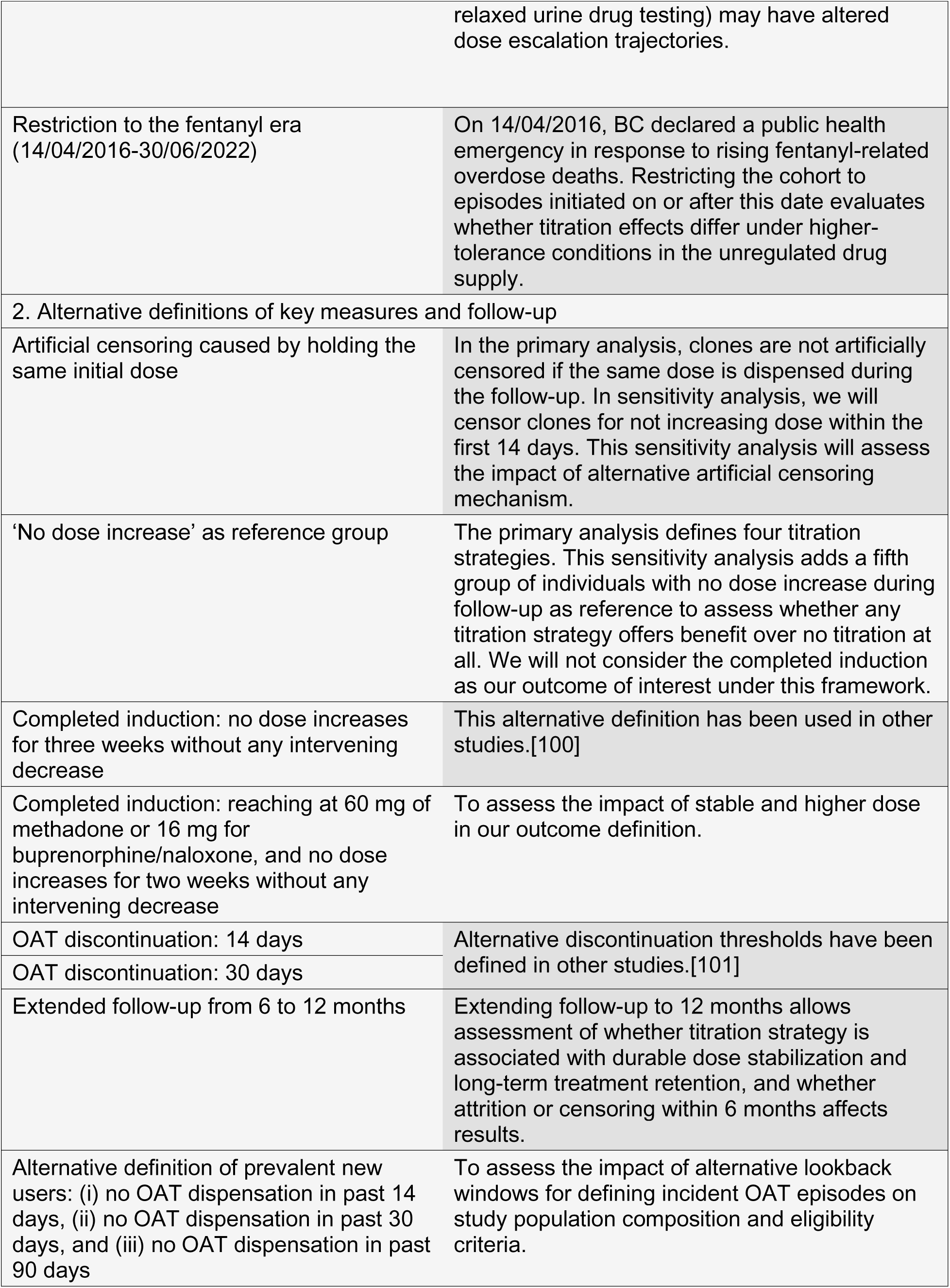
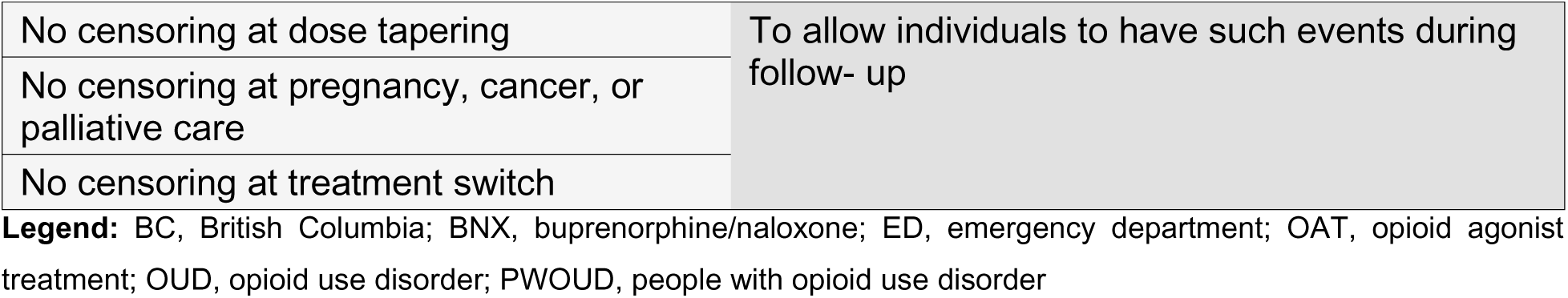
Proposed subgroup and sensitivity analyses.

### Ethics and Dissemination

The linked databases are made available to the research team by the British Columbia Ministries of Health and of Mental Health and Addictions as part of the response to the provincial opioid overdose public health emergency. The data are fully de-identified before being provided to the research team, and no identifiable personal information was accessed. The study is classified as a quality improvement initiative. The Providence Health Care Research Institute and the Simon Fraser University Office of Research Ethics determine that the analysis meet the criteria for exemption per Article 2.5 of the Tri-Council Policy Statement: Ethical Conduct for Research Involving Humans. Since the study uses de-identified administrative data and involves no direct contact with individuals, the requirement for informed consent is waived by the reviewing bodies.

This study will adhere to the Transparent Reporting of Observational Studies Emulating a Target Trial (the TARGET statement) for conducting and reporting research. Results will be disseminated to clinicians, local advocacy groups and decision-makers, and national and international clinical guideline committees, presented at international conferences, and published in peer-reviewed journals both electronically and in print.

### Study status and timeline

The de-identified administrative health data covering the study period (01/01/2010 to 31/12/2022) have been obtained, but no analyses have been conducted to date. Data cleaning, cohort construction, and all planned analyses will begin following protocol approval and the study is expected to be completed within eight months afterwards.

## Discussion

This study will generate population-level evidence on the comparative effectiveness of alternative dose titration schedules for methadone and buprenorphine/naloxone in the treatment of OUD. Current guidelines recommend scheduled titration based largely on expert consensus, with limited real-world evidence on how quickly dose should be titrated, a gap of particular consequence amid widespread fentanyl exposure. By emulating per-protocol target trials with linked administrative data, we will estimate the effect of titration schedules ranging from the most conservative to the most aggressive permissible pace on completed induction and all-cause mortality, using a CCW approach.

A key limitation is that several determinants of dosing decisions, OUD severity, in-hospital OAT dispensing, and evolving clinical circumstances prompting dose changes, are incompletely captured in administrative data. Since our strategies reflect the maximum permissible titration pace rather than a single prescribed schedule, the implications of these definitional choices are examined across several sensitivity analyses. Our increment-by-interval framework also does not uniquely identify rapid in-hospital or low-dose (‘micro-dosing’) inductions: the former occurs during hospitalisation, where inpatient dosing is not captured in our data, while the latter involves within-day or sub-therapeutic dose escalation that daily dispensing records cannot resolve. These practices, though highlighted in the Introduction, are therefore not directly compared here. In addition, dispensing records confirm medication availability but not ingestion; exposure misclassification is therefore possible if dispensed doses are not consumed as directed. Despite these constraints, the large population coverage, rigorous target-trial design, and extensive sensitivity analyses will provide robust, policy-relevant evidence to inform titration guidance for methadone and buprenorphine/naloxone in jurisdictions facing escalating opioid-related challenges.

### Patient and public involvement

Although no patients were directly engaged in designing this study, its conceptualization was shaped by previous interactions with local advocacy organizations representing individuals who use drugs and those accessing OAT.[102] Qualitative feedback on this study and other related objectives outlined in the parent grant R01DA050629 was incorporated to prioritize this analysis, taking into account its potential impact on completed induction and client engagement. The results will be disseminated to local advocacy groups after the analysis is completed.

## Data Availability

Access to data provided by the Data Stewards is subject to approval and may be requested for research projects through the Data Stewards or their designated service providers. The data used in this study are not publicly available because of data-sharing agreements and privacy restrictions.

## Acknowledgements

Contributors: Mondol conducted literature reviews, conceived the design and analysis strategy, wrote the first draft of the article. Zanette conducted literature reviews and critical revisions. Nosyk, Min, Zanette, Kurz, Platt, Seaman, Gustafson, Socías, and Sutherland provided critical revisions. Nosyk conceptualized and secured funding for the study. All authors approved the final draft.

## Disclaimer

Access to data provided by the Data Stewards is subject to approval but can be requested for research projects through the Data Stewards or their designated service providers. The following data sets were used in this study: PharmaNet, Client Roster, Medical Services Plan, Discharge Abstract Database, National Ambulatory Care Reporting System Database, Provincial Corrections, Perinatal Database, Social Development and Poverty Reduction Database, and Vital Statistics. All inferences, opinions, and conclusions drawn in this publication are those of the author(s), and do not reflect the opinions or policies of the Data Steward(s). This Data was provisioned under ISP 17-162.

## Role of the Funding source

This study was funded by the National Institutes on Drug Abuse (NIDA grant no. R01DA050629). The funding source was independent of the design of this study and did not have any role during its execution, analyses, interpretation of the data, writing or decision to submit results. The authors had full access to the results in the study and took responsibility for the integrity of the data and accuracy of the analysis.

## Notes

### Competing Interest Statement

The authors have declared no competing interest.

### Author Declarations

Ethics committee/IRB of the Providence Health Care Research Institute and the Simon Fraser University Office of Research Ethics waived ethical approval for this work. This study will use fully de-identified administrative health databases from British Columbia, Canada and was classified as a quality improvement initiative. The reviewing bodies determined that the study meets the criteria for exemption under Article 2.5 of the Tri-Council Policy Statement: Ethical Conduct for Research Involving Humans.

