## Supplementary material for "What constitutes safe and effective dose titration of methadone and buprenorphine/naloxone? protocol for a population-based target trial emulation": S1 Table, S2 Table, S1 Fig.

### Supplementary Tables

#### S1 Table. Drug identification numbers for identification of opioid agonist treatment from PharmaNet

| OAT | DIN/PIN* |
| --- | --- |
| Methadone | 999792, 999793, 66999990, 66999991, 66999992, 66999993, 66999997, 66999998, 66999999, 67000000, 67000001, 67000002, 67000003, 67000004, 67000005, 67000006, 67000007, 67000008, 67000009,67000010, 67000011, 67000012, 67000013, 67000014, 67000015, 67000016, 67000017, 67000018, 67000019, 67000020 |
| Buprenorphine/naloxone | 2295695, 2295709, 2408090, 2408104, 2424851, 2424878, 2453908, 2453916, 2468085, 2468093, 2502313, 2502321, 2502348, 2502356, 2517175, 2517183 |
| Slow-release oral morphine (Kadian) | 22123349, 22123346, 22123347, 22123348 |
| Injectable OAT^†^ | 2146126, 22123340, 22123357, 2469413, 66123367 |
| t-IOAT (Hydromorphone) | 786543, 885428 |

*Abbreviations:* OAT: opioid agonist treatment. *Drug Identification Numbers (DIN)/Product Identification Numbers (PIN); ^†^Diacetylmorphine or hydromorphone with some restrictions based on prescriber, dispensing pharmacy and/or date.

#### S2 Table. List of linked administrative health data sources

| Database | Description |
| --- | --- |
| Medical Services Plan (MSP) [1] | All medically necessary services provided by fee-for-service practitioners covered by the province’s universal insurance program. |
| Discharge Abstract Database (DAD) [2] | All hospital discharges, day surgery, transfers and deaths of inpatients, data of BC residents treated at hospital out of province and out-of-province residents treated within BC hospitals included. |
| PharmaNet (PNET) [3] | All prescriptions for drugs and medical supplies dispensed from community pharmacies. |
| BC Vital Statistics (BCVS) [4] | All death and their underlying causes registered in the province. |
| BC Corrections [5] | Date of intake into and discharge from the correctional system, discharge location and reason for release. |
| National Ambulatory Care Reporting System database (NACRS) [6] | All hospital-based and community-based ambulatory care including day surgery, outpatient and community-based clinics emergency departments. |
| BC Perinatal database (BCPD) [7] | Maternal and child neonatal health for all provincial births data collected from obstetrical facilities as well as births occurring at home attended by BC Registered Midwives. |
| Client roster databases[8] | Demographic and geographic information for the Ministry’s clients. |
| Social Development and Poverty Reduction (SDPR) [9] | Persons who received income or disability assistance by the ministry of SDPR; the data contain type of assistance program, family type, a number of dependents, payment and homelessness flag. |
| MSP, DAD, PNET, BCVS, and client roster databases are available from 01/01/1996 to 12/31/2022. BC Corrections and SDPR are available from 01/01/2010 to 12/31/2022. The NACRS available from 04/01/2012 to 12/31/2022. The BCPD is available from 08/01/2000 to 12/31/2022. The study period begins 01/01/2010, and ends 30/06/2022. | |

### Supplementary Figures

#### S1 Fig: Construction of OAT episodes.

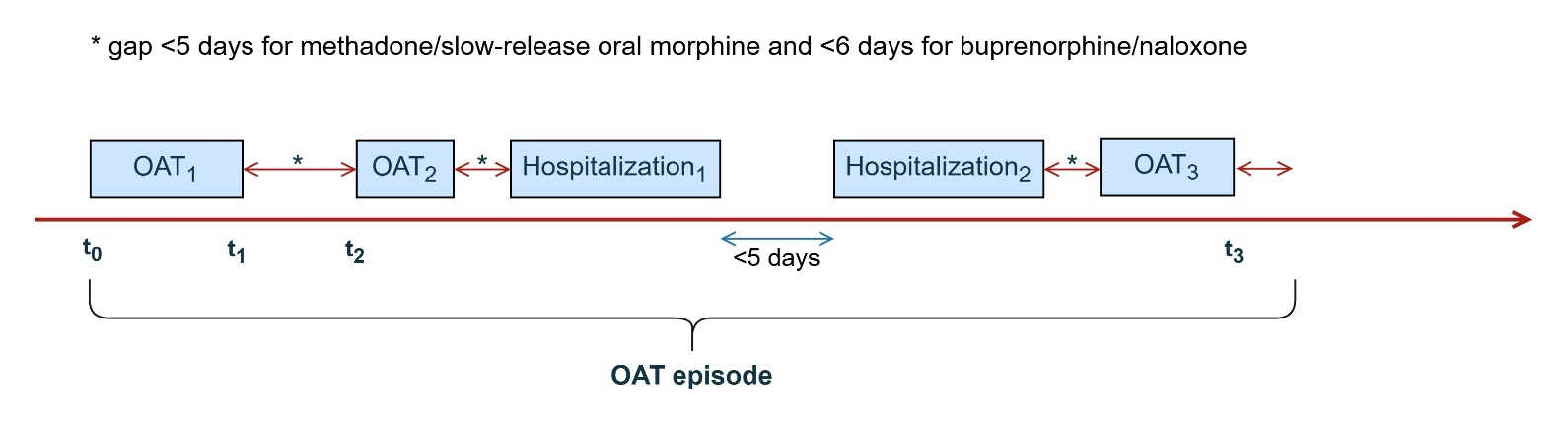

**Legend:** OAT episode is defined as continuous dispensation without having an interruption of ≥5 days for methadone/slow-release oral morphine, ≥6 days for buprenorphine/naloxone, and ≥3 days for injectable OAT. The definition follows clinical guidelines for reversion to initial dosing after these interruptions. Individuals on OAT continue their doses while in the hospital. The episode length [t_3_ - t_1_ + 5] for methadone and [t_3_ - t_1_ + 6] for buprenorphine/naloxone will be considered for OAT discontinuation outcome.
